# The effect of coronary revascularization timing on cardiovascular outcomes in patients with ischemic heart disease

**DOI:** 10.1101/2024.06.12.24308844

**Authors:** Sean Hardiman, Guy Fradet, Lisa Kuramoto, Michael Law, Simon Robinson, Boris Sobolev

**Affiliations:** School of Population and Public Health, University of British Columbia, Vancouver, Canada; Division of Cardiovascular Surgery, Department of Surgery, Faculty of Medicine, University of British Columbia, Vancouver, Canada; Centre for Clinical Epidemiology and Evaluation, Vancouver Coastal Health Research Institute, Vancouver, Canada; Division of Cardiology, Department of Medicine, Faculty of Medicine, University of British Columbia, Vancouver, Canada

**Keywords:** Comparative effectiveness, Coronary artery bypass graft, Percutaneous coronary intervention, Cardiovascular outcomes, Treatment timing

## Abstract

**Background:** There is little evidence on whether the timing of revascularization affects cardiovascular disease progression in patients with stable ischemic heart disease. We aimed to determine if disease progression differed between patients who underwent coronary artery bypass graft (CABG) surgery after the time recommended by physicians compared to timely percutaneous coronary intervention (PCI).

**Methods:** We identified 25,469 British Columbia, Canada residents ages 60 years or older who underwent their first non-emergency revascularization for angiographically proven, stable left main or multi-vessel ischemic heart disease. We estimated the cumulative incidence of a composite cardiovascular outcome (CVO) and death as a competing risk for patients undergoing delayed CABG versus timely PCI.

**Results:** After adjustment, patients who underwent delayed CABG had a statistically significantly lower cumulative CVO incidence at three years compared with those who received timely PCI (9.6% delayed CABG, 23.2% timely PCI; subdistribution hazard ratio for CVO at three years 0.50, 95% CI 0.26–0.99).

**Conclusion:** Our results suggest that for patients who wish to wait for CABG, doing so may lead to slower disease progression compared with receiving PCI.

## INTRODUCTION

In Canada, multiple factors contribute to the timing of coronary artery disease treatment, including clinical need, demand variation, and budget allocations.^1^ Thus, patients with stable ischemic heart disease where non-emergency coronary revascularization is required, either by coronary artery bypass graft (CABG) surgery or percutaneous coronary intervention (PCI), may experience delays during times of greater demand or reduced supply.^2^

Prior research has shown that patients waiting for CABG benefit from lower mortality when offered earlier timing of treatment.^3^ However, no research is available to show if a similar benefit in cardiovascular disease progression exists. Patients with multi-vessel or left-main coronary artery disease who do not need emergency treatment should consider CABG rather than PCI,^4^ due to lower mortality in some populations, fewer post-procedural acute myocardial infarctions (AMI), and a reduced need for repeat revascularization. These recommendations are based primarily on the randomized controlled trial evidence in patients with stable multi-vessel disease and left main disease. These trials have had varying primary outcomes, with most using some variation of Major Adverse Cardiovascular Events (MACE), a composite outcome that includes death and other disease or procedure-related outcomes. The choice of composite outcomes has been criticized in the CABG versus PCI literature,^5^ due to the high rate of repeat revascularization observed in patients that undergo PCI that contribute to statistically significant differences where otherwise, none might be observed.

While randomized trials have included some elements of cardiovascular disease progression in their composite outcomes, none have used a disease progression construct or included patients with delays in treatment. Therefore, we established our primary research question: does the cumulative incidence of cardiovascular disease progression differ in patients with stable multi-vessel or left main ischemic heart disease after delayed CABG compared to timely PCI?

Our paper has two objectives: Estimate the cumulative incidence of composite CVO, in the presence of death as a competing risk, in patients with stable multi-vessel or left main ischemic heart disease after CABG with delay and PCI within appropriate time (1) without adjustment and (2) adjusting for patient, disease, and treatment characteristics.

## STUDY DATA AND METHODS

This study follows the Strengthening the Reporting of Observational Studies in Epidemiology (STROBE) guidelines for the reporting of observational cohort studies.^6^

### Study design

We conducted a retrospective cohort study of prospectively collected data amongst all patients in the province of BC who underwent isolated CABG surgery or PCI for the treatment of coronary artery disease.

### Data sources

We obtained diagnostic catheterization, PCI, and isolated CABG surgery records from the provincial registries maintained by Cardiac Services BC (CSBC), a program of the Provincial Health Services Authority (Vancouver, BC, Canada). We used CSBC’s diagnostic catheterization, CABG, and PCI registry data to construct an episode of care, which contains all events occurring from diagnostic catheterization through to revascularization. We linked these care episodes to the BC Ministry of Health’s Discharge Abstract Database (DAD), which contains hospitalization records, and the BC Vital Statistics Deaths file, which contains mortality data. Finally, we linked this data set to Population Data BC’s Central Demographics File, which contains demographic data for all study participants.

### Setting & Participants

The study cohort includes patients aged 60 years or older, who underwent non-emergency first-time revascularization for angiographically-proven, stable left main or multi-vessel ischemic heart disease in BC, between January 1, 2001, and December 31, 2016. We defined revascularization as either a PCI or an isolated CABG surgery. Patient age, extent of disease, and non-emergency status were identified using the Cardiac Services BC cardiac surgery and PCI registry data. Stable disease was identified using atherosclerotic heart disease code (ICD-10-CA I25.0, I25.1, I25.10; ICD-9 429.2 414.0) logged as diagnosis type M (most responsible), type 1 (pre-admit comorbidity), type 2 (post-admit comorbidity), type 6 (proxy most responsible diagnosis), or types W, X, or Y (first, second, or third service transfers) in the DAD. The index event in this study is first-ever revascularization, by either PCI or CABG, within the study period of January 1, 2001, and December 31, 2016.

### Variables

#### Study variable

The study variable is treatment timing, operationalized as the time to coronary revascularization treatment and computed in calendar days. The time to treatment starts on the date when the need for revascularization is clinically established and the patient is ready, willing, and able to undergo revascularization, operationalized as the booking date in CSBC records. The time to treatment ends on the date the revascularization procedure was performed. Dates to calculate treatment timing were collected from CSBC registries for CABG and PCI. Patients were assigned to one of two study groups: delayed CABG or timely PCI. Timing groups were established based on the First Minsters’ Meeting benchmarks^7^ in use by CSBC to prioritize patients for CABG and the Canadian Cardiovascular Society (CCS) Access to Care recommendations^8^ for CABG and PCI (Table 1).

**Table 1.** Study groups by procedure type and urgency and treatment delay in days.

| Procedure | CSBC Urgency | Timely Treatment |  | Delayed Treatment |
| --- | --- | --- | --- | --- |
|  |  | Interval Start | Interval End | Interval Start |
| CABG | Priority I | 1 Day | 7 Days | 8+ Days |
|  | Priority II | 1 Day | 14 Days | 15+ Days |
|  | Priority III | 1 Day | 42 Days | 43+ Days |
| PCI | Urgent Inpatient | 1 Day | 5 Days | 6+ Days |
|  | Urgent Outpatient | 1 Day | 14 Days | 15+ Days |
|  | Elective | 1 Day | 42 Days | 43+ Days |

#### Outcome variable

The outcome variable is the time to cardiovascular disease progression, operationalized as the first cardiovascular outcome (CVO), a composite outcome. The composite outcome records the first hospital admission after index revascularization or, in the case of patients with multiple PCI records, after the last staged PCI, for either AMI, unstable angina, congestive heart failure, or cerebrovascular accident (CVA). We identified the occurrence of each outcome using ICD-9 or ICD-10-CA diagnosis codes found in the DAD with diagnosis types ‘M’, ‘1’, ‘2’, ‘6’, ‘W’, ‘X’, or ‘Y’. We operationalized components of the composite outcome starting with ICD-10-CA codes: AMI, I21 and I22; unstable angina, I20.0; and CHF, I50. CVA was operationalized using ICD-10-CA codes from the Stroke Council of the American Heart Association’s definition of Stroke (Supplementary Material).^9,10^ Equivalent ICD-9 codes for all components of the composite were established through application of the Canadian Institute for Health Information (CIHI) conversion tables^11^ to the ICD-10-CA codes. We used the date of death from any cause recorded in the BC Vital Statistics Deaths File. We measured the time to event from either the index revascularization for CABG and single-session PCI or the last staged PCI in patients with multiple PCI records until a CVO, death, three years’ follow-up, or study end. Due to data collection gaps in staged PCI data available from CSBC, we established an algorithm to identify the last staged PCI using published guidance,^12^ based on a common diagnostic catheterization date to the index revascularization. We also developed rules to differentiate last staged PCI and repeat revascularization in patients with multiple PCI records (Supplementary Material).

### Statistical Methods

We estimated the frequency and percentage of patients by characteristics and by treatment group. Groups were compared using a chi-square test for categorical variables and p-values for between group differences reported. We modelled the cumulative incidence function (CIF) of CVO in the presence of death as a competing risk, for each treatment group over three years using with a flexible parametric approach using restricted cubic spline functions.^13^ We reported the unadjusted cumulative incidence at three years, and the unadjusted subdistribution hazard ratio of CVO, in the presence of death as a competing risk, at three years, for each study group.^14^ The subdistribution hazard ratio for CVO gives the association between treatment received and the CVO-specific CIF. A subdistribution hazard ratio of less than one means the delayed CABG group had a lower subdistribution hazard of a composite cardiovascular outcome at 3 years compared to the timely PCI group, in the presence of death as a competing risk. A hazard greater than one means the delayed CABG group had a higher subdistribution hazard of a composite cardiovascular outcome at 3 years compared to the timely PCI group, in the presence of death as a competing risk.

We then estimated propensity scores^15^ for the probability of belonging to each study group using logistic regression and, using those scores, calculated inverse probability of treatment weights.^16^ Each patient was weighted by the inverse of the probability of being assigned to their treatment group to adjust for differences between the two treatment groups. We assessed the performance of the propensity score model by comparing the distribution of covariates and propensity scores before and after inverse probability weighting and by calculating standardized mean differences. Adjusted CIFs and subdistribution hazard ratios at three years were obtained using an inverse probability weighted flexible parametric approach. Statistical analyses were performed using Stata 17 (College Station, TX). Flexible parametric models were constructed using *stpm2cr*, a Stata software package.^17^

### Patient and public involvement

We consulted the Pacific Open-Heart Association (POHA), a cardiac surgery peer support group in Vancouver, BC, to inform our research query. They confirmed that patients frequently wait for CABG and that the wait contributes to anxiety, given that a planned heart surgery could be scheduled at any moment. In this study, we answer the question that patients told us matters most: is it be better for CABG candidates to undergo PCI instead of waiting an unknown time for CABG?

## RESULTS

### Participants

We identified 39,176 patients who met the selection criteria for our study (Figure 1). We did not select patients for the analytical cohort if their revascularization record could not be linked to hospital records (*n*=556), their PCI record was for ad-hoc PCI, but the procedure could not be linked to a diagnostic catheterization (*n*=35), or that their hospital records did not contain diagnosis codes indicative of stable ischemic heart disease (*n*=696). 37,889 patients were eligible for analysis.

**Figure 1.**
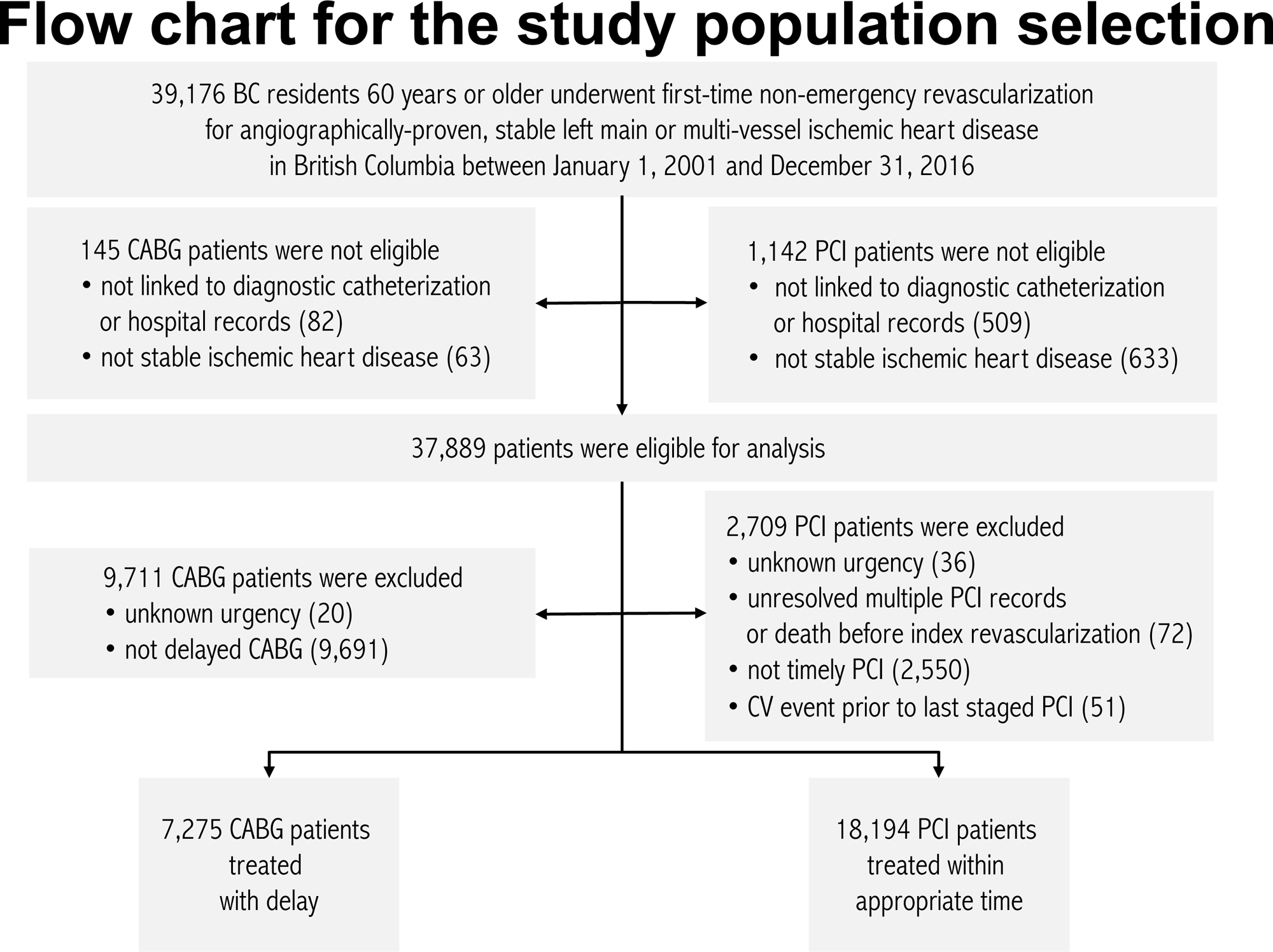
Flow diagram for the study population selection.

We set aside patients if their procedure urgency could not be determined (*n*=56), if patients with multiple PCI records were unresolved or if there were errors in the administrative data set where date of death preceded date of revascularization (*n*=72), if patients had a CVO between index revascularization and the last staged PCI (*n*=51), if the patient received delayed PCI (*n*=2,550), or if the patient received timely CABG (*n*=9,711). 25,469 patients were available to be analyzed.

### Descriptive data

The baseline characteristics of the patients in the analytical cohort are shown in Table 2.

**Table 2.**
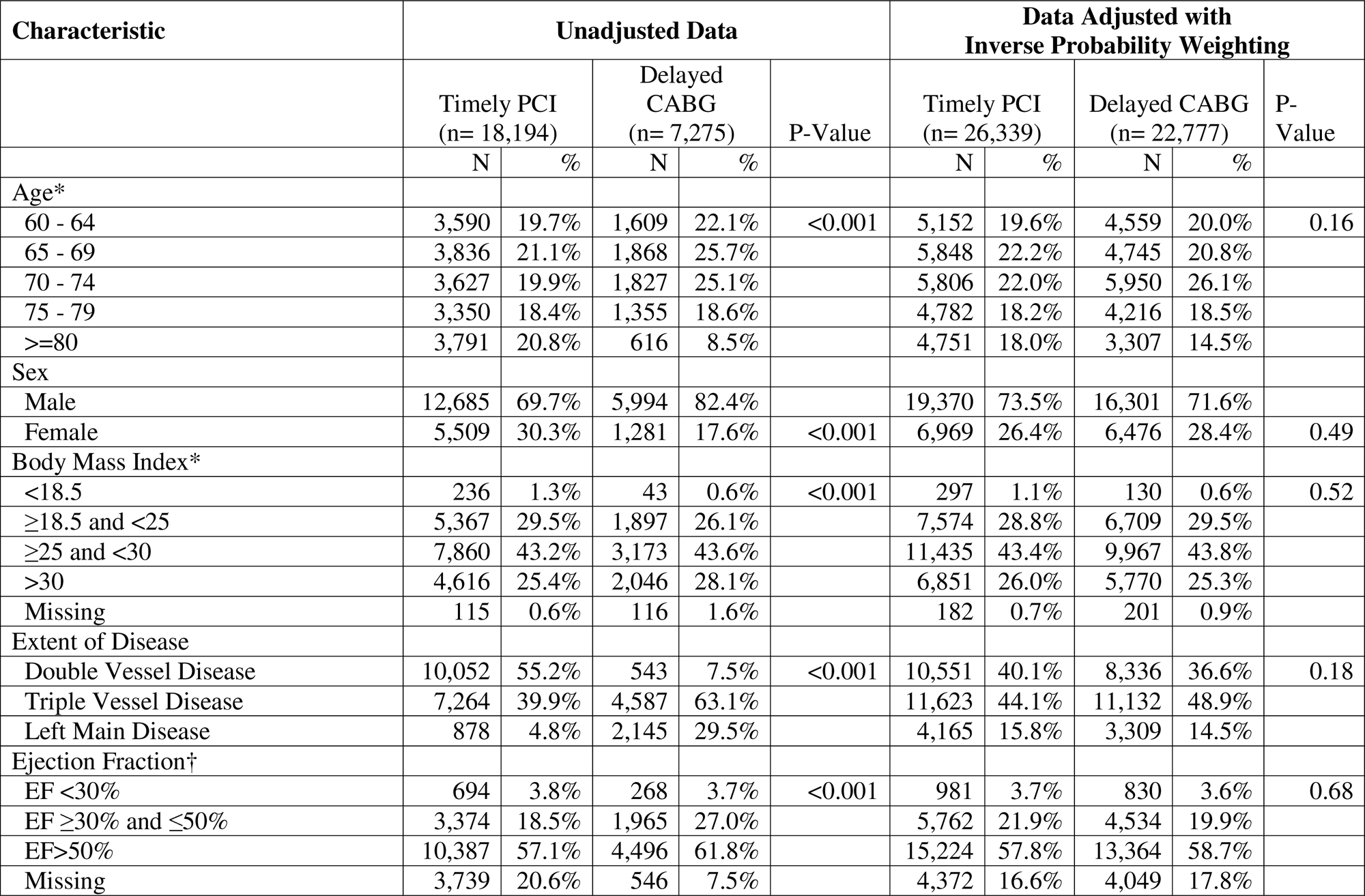
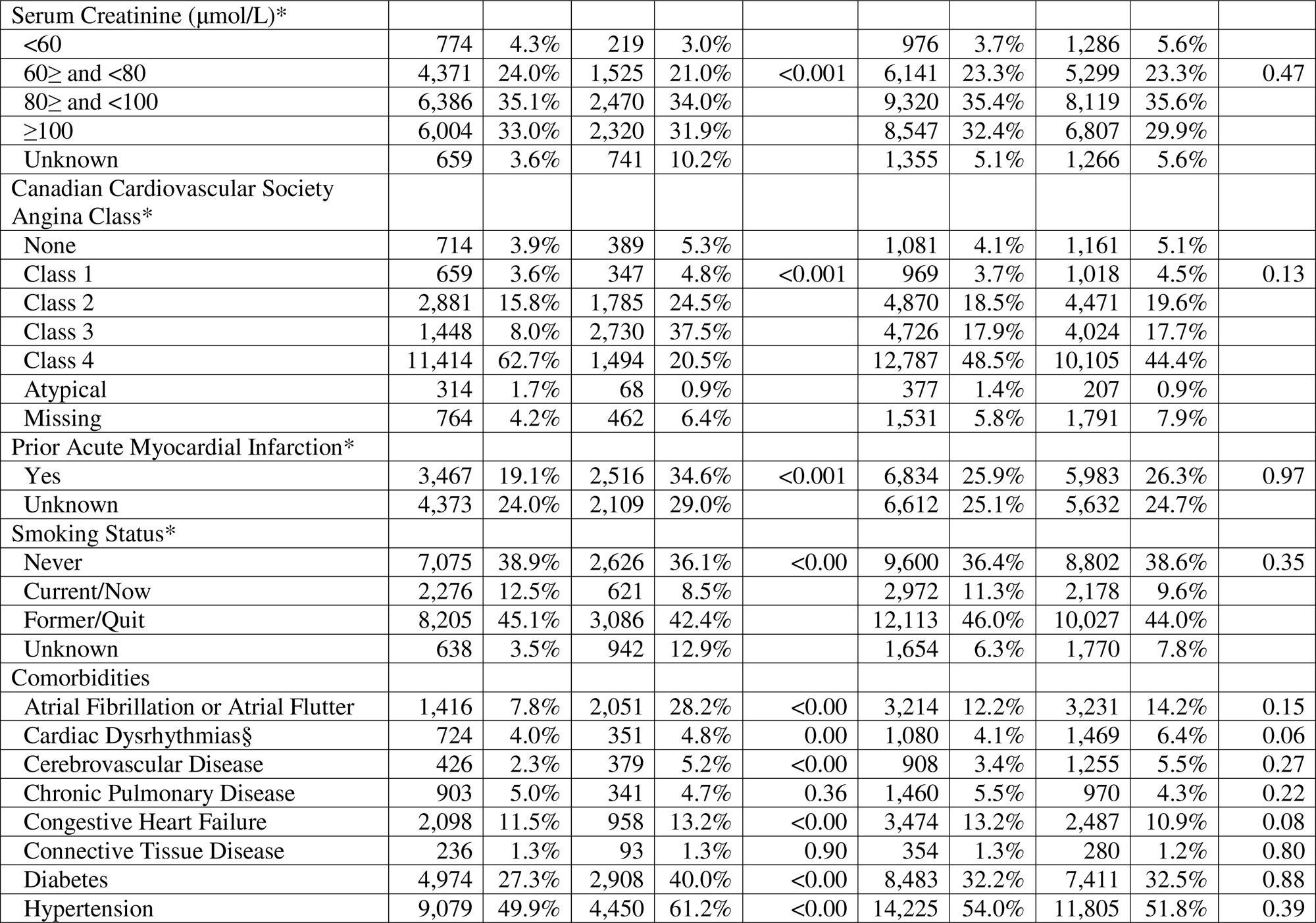
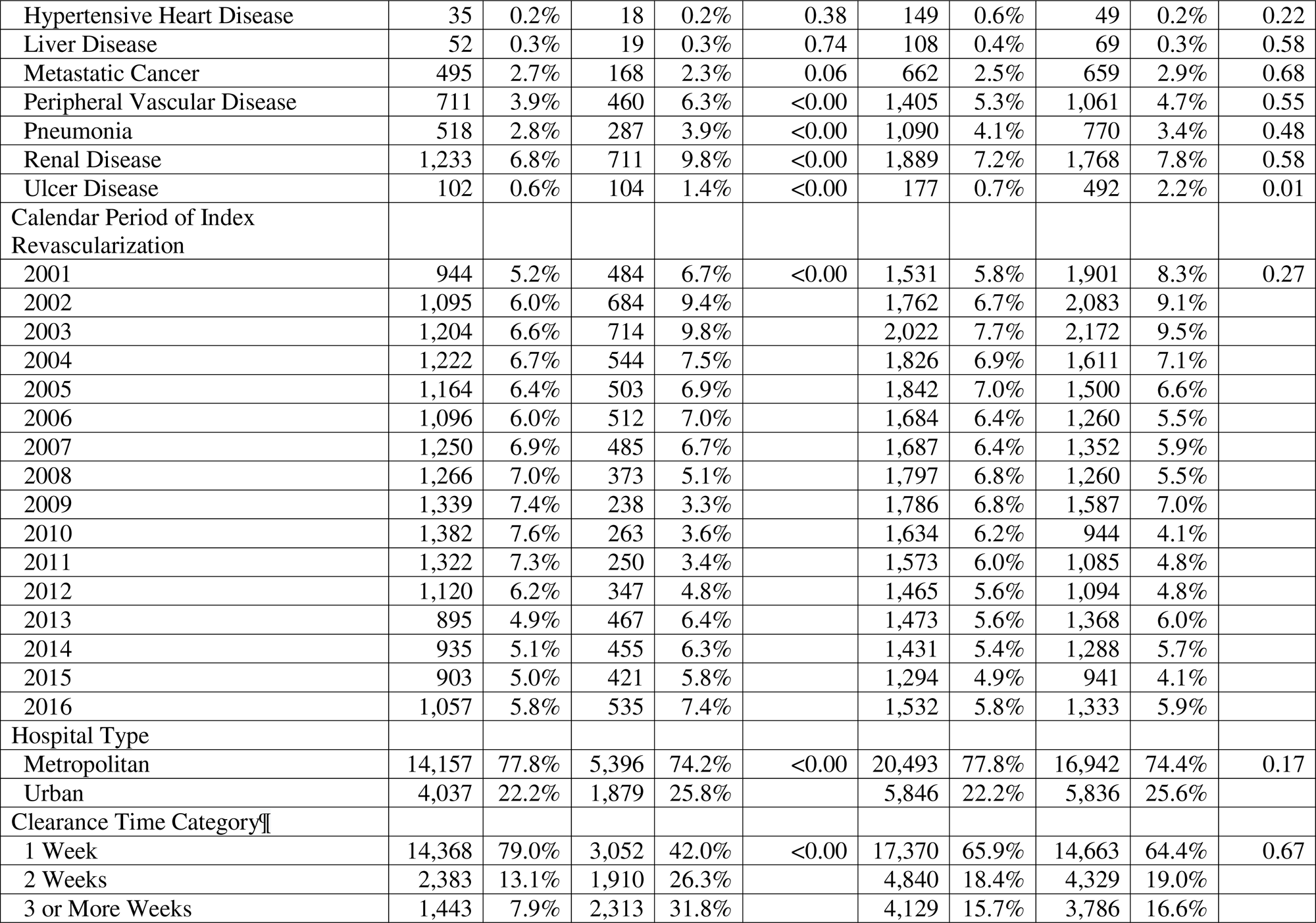
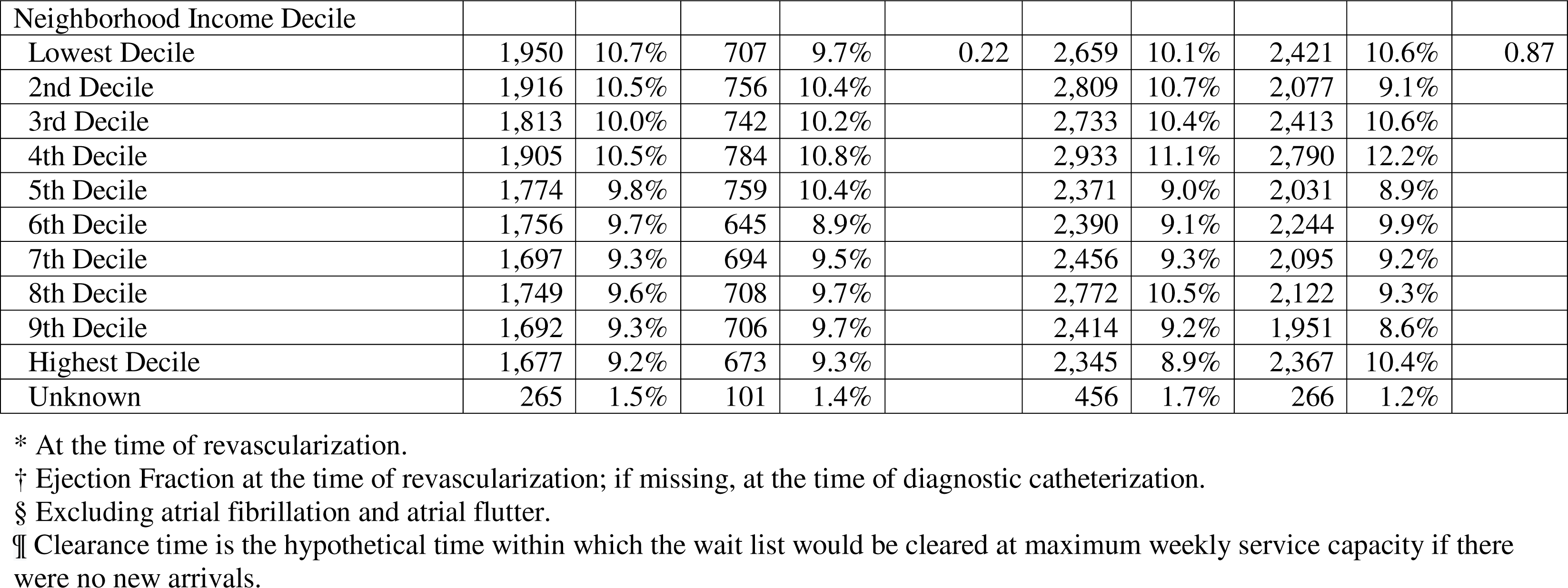
Baseline characteristics of the patients.

| Characteristic | Unadjusted Data |  |  |  |  | Data Adjusted with<br>Inverse Probability Weighting |  |  |  |  |
| --- | --- | --- | --- | --- | --- | --- | --- | --- | --- | --- |
|  | Timely PCI<br>(n= 18,194) |  | Delayed<br>CABG<br>(n= 7,275) |  | P-Value | Timely PCI<br>(n= 26,339) |  | Delayed CABG<br>(n= 22,777) |  | P-Value |
|  | N | % | N | % |  | N | % | N | % |  |
| Age* |  |  |  |  |  |  |  |  |  |  |
| 60 - 64 | 3,590 | 19.7% | 1,609 | 22.1% | <0.001 | 5,152 | 19.6% | 4,559 | 20.0% | 0.16 |
| 65 - 69 | 3,836 | 21.1% | 1,868 | 25.7% |  | 5,848 | 22.2% | 4,745 | 20.8% |  |
| 70 - 74 | 3,627 | 19.9% | 1,827 | 25.1% |  | 5,806 | 22.0% | 5,950 | 26.1% |  |
| 75 - 79 | 3,350 | 18.4% | 1,355 | 18.6% |  | 4,782 | 18.2% | 4,216 | 18.5% |  |
| ≥80 | 3,791 | 20.8% | 616 | 8.5% |  | 4,751 | 18.0% | 3,307 | 14.5% |  |
| Sex |  |  |  |  |  |  |  |  |  |  |
| Male | 12,685 | 69.7% | 5,994 | 82.4% |  | 19,370 | 73.5% | 16,301 | 71.6% |  |
| Female | 5,509 | 30.3% | 1,281 | 17.6% | <0.001 | 6,969 | 26.4% | 6,476 | 28.4% | 0.49 |
| Body Mass Index* |  |  |  |  |  |  |  |  |  |  |
| <18.5 | 236 | 1.3% | 43 | 0.6% | <0.001 | 297 | 1.1% | 130 | 0.6% | 0.52 |
| ≥18.5 and <25 | 5,367 | 29.5% | 1,897 | 26.1% |  | 7,574 | 28.8% | 6,709 | 29.5% |  |
| ≥25 and <30 | 7,860 | 43.2% | 3,173 | 43.6% |  | 11,435 | 43.4% | 9,967 | 43.8% |  |
| >30 | 4,616 | 25.4% | 2,046 | 28.1% |  | 6,851 | 26.0% | 5,770 | 25.3% |  |
| Missing | 115 | 0.6% | 116 | 1.6% |  | 182 | 0.7% | 201 | 0.9% |  |
| Extent of Disease |  |  |  |  |  |  |  |  |  |  |
| Double Vessel Disease | 10,052 | 55.2% | 543 | 7.5% | <0.001 | 10,551 | 40.1% | 8,336 | 36.6% | 0.18 |
| Triple Vessel Disease | 7,264 | 39.9% | 4,587 | 63.1% |  | 11,623 | 44.1% | 11,132 | 48.9% |  |
| Left Main Disease | 878 | 4.8% | 2,145 | 29.5% |  | 4,165 | 15.8% | 3,309 | 14.5% |  |
| Ejection Fraction† |  |  |  |  |  |  |  |  |  |  |
| EF <30% | 694 | 3.8% | 268 | 3.7% | <0.001 | 981 | 3.7% | 830 | 3.6% | 0.68 |
| EF ≥30% and ≤50% | 3,374 | 18.5% | 1,965 | 27.0% |  | 5,762 | 21.9% | 4,534 | 19.9% |  |
| EF>50% | 10,387 | 57.1% | 4,496 | 61.8% |  | 15,224 | 57.8% | 13,364 | 58.7% |  |
| Missing | 3,739 | 20.6% | 546 | 7.5% |  | 4,372 | 16.6% | 4,049 | 17.8% |  |
| Serum Creatinine (μmol/L)* |  |  |  |  |  |  |  |  |  |  |
| <60 | 774 | 4.3% | 219 | 3.0% |  | 976 | 3.7% | 1,286 | 5.6% |  |
| 60≥ and <80 | 4,371 | 24.0% | 1,525 | 21.0% | <0.001 | 6,141 | 23.3% | 5,299 | 23.3% | 0.47 |
| 80≥ and <100 | 6,386 | 35.1% | 2,470 | 34.0% |  | 9,320 | 35.4% | 8,119 | 35.6% |  |
| ≥100 | 6,004 | 33.0% | 2,320 | 31.9% |  | 8,547 | 32.4% | 6,807 | 29.9% |  |
| Unknown | 659 | 3.6% | 741 | 10.2% |  | 1,355 | 5.1% | 1,266 | 5.6% |  |
| Canadian Cardiovascular Society Angina Class* |  |  |  |  |  |  |  |  |  |  |
| None | 714 | 3.9% | 389 | 5.3% |  | 1,081 | 4.1% | 1,161 | 5.1% |  |
| Class 1 | 659 | 3.6% | 347 | 4.8% | <0.001 | 969 | 3.7% | 1,018 | 4.5% | 0.13 |
| Class 2 | 2,881 | 15.8% | 1,785 | 24.5% |  | 4,870 | 18.5% | 4,471 | 19.6% |  |
| Class 3 | 1,448 | 8.0% | 2,730 | 37.5% |  | 4,726 | 17.9% | 4,024 | 17.7% |  |
| Class 4 | 11,414 | 62.7% | 1,494 | 20.5% |  | 12,787 | 48.5% | 10,105 | 44.4% |  |
| Atypical | 314 | 1.7% | 68 | 0.9% |  | 377 | 1.4% | 207 | 0.9% |  |
| Missing | 764 | 4.2% | 462 | 6.4% |  | 1,531 | 5.8% | 1,791 | 7.9% |  |
| Prior Acute Myocardial Infarction* |  |  |  |  |  |  |  |  |  |  |
| Yes | 3,467 | 19.1% | 2,516 | 34.6% | <0.001 | 6,834 | 25.9% | 5,983 | 26.3% | 0.97 |
| Unknown | 4,373 | 24.0% | 2,109 | 29.0% |  | 6,612 | 25.1% | 5,632 | 24.7% |  |
| Smoking Status* |  |  |  |  |  |  |  |  |  |  |
| Never | 7,075 | 38.9% | 2,626 | 36.1% | <0.00 | 9,600 | 36.4% | 8,802 | 38.6% | 0.35 |
| Current/Now | 2,276 | 12.5% | 621 | 8.5% |  | 2,972 | 11.3% | 2,178 | 9.6% |  |
| Former/Quit | 8,205 | 45.1% | 3,086 | 42.4% |  | 12,113 | 46.0% | 10,027 | 44.0% |  |
| Unknown | 638 | 3.5% | 942 | 12.9% |  | 1,654 | 6.3% | 1,770 | 7.8% |  |
| Comorbidities |  |  |  |  |  |  |  |  |  |  |
| Atrial Fibrillation or Atrial Flutter | 1,416 | 7.8% | 2,051 | 28.2% | <0.00 | 3,214 | 12.2% | 3,231 | 14.2% | 0.15 |
| Cardiac Dysrhythmias§ | 724 | 4.0% | 351 | 4.8% | 0.00 | 1,080 | 4.1% | 1,469 | 6.4% | 0.06 |
| Cerebrovascular Disease | 426 | 2.3% | 379 | 5.2% | <0.00 | 908 | 3.4% | 1,255 | 5.5% | 0.27 |
| Chronic Pulmonary Disease | 903 | 5.0% | 341 | 4.7% | 0.36 | 1,460 | 5.5% | 970 | 4.3% | 0.22 |
| Congestive Heart Failure | 2,098 | 11.5% | 958 | 13.2% | <0.00 | 3,474 | 13.2% | 2,487 | 10.9% | 0.08 |
| Connective Tissue Disease | 236 | 1.3% | 93 | 1.3% | 0.90 | 354 | 1.3% | 280 | 1.2% | 0.80 |
| Diabetes | 4,974 | 27.3% | 2,908 | 40.0% | <0.00 | 8,483 | 32.2% | 7,411 | 32.5% | 0.88 |
| Hypertension | 9,079 | 49.9% | 4,450 | 61.2% | <0.00 | 14,225 | 54.0% | 11,805 | 51.8% | 0.39 |
| Hypertensive Heart Disease | 35 | 0.2% | 18 | 0.2% | 0.38 | 149 | 0.6% | 49 | 0.2% | 0.22 |
| Liver Disease | 52 | 0.3% | 19 | 0.3% | 0.74 | 108 | 0.4% | 69 | 0.3% | 0.58 |
| Metastatic Cancer | 495 | 2.7% | 168 | 2.3% | 0.06 | 662 | 2.5% | 659 | 2.9% | 0.68 |
| Peripheral Vascular Disease | 711 | 3.9% | 460 | 6.3% | <0.00 | 1,405 | 5.3% | 1,061 | 4.7% | 0.55 |
| Pneumonia | 518 | 2.8% | 287 | 3.9% | <0.00 | 1,090 | 4.1% | 770 | 3.4% | 0.48 |
| Renal Disease | 1,233 | 6.8% | 711 | 9.8% | <0.00 | 1,889 | 7.2% | 1,768 | 7.8% | 0.58 |
| Ulcer Disease | 102 | 0.6% | 104 | 1.4% | <0.00 | 177 | 0.7% | 492 | 2.2% | 0.01 |
| Calendar Period of Index<br>Revascularization |  |  |  |  |  |  |  |  |  |  |
| 2001 | 944 | 5.2% | 484 | 6.7% | <0.00 | 1,531 | 5.8% | 1,901 | 8.3% | 0.27 |
| 2002 | 1,095 | 6.0% | 684 | 9.4% |  | 1,762 | 6.7% | 2,083 | 9.1% |  |
| 2003 | 1,204 | 6.6% | 714 | 9.8% |  | 2,022 | 7.7% | 2,172 | 9.5% |  |
| 2004 | 1,222 | 6.7% | 544 | 7.5% |  | 1,826 | 6.9% | 1,611 | 7.1% |  |
| 2005 | 1,164 | 6.4% | 503 | 6.9% |  | 1,842 | 7.0% | 1,500 | 6.6% |  |
| 2006 | 1,096 | 6.0% | 512 | 7.0% |  | 1,684 | 6.4% | 1,260 | 5.5% |  |
| 2007 | 1,250 | 6.9% | 485 | 6.7% |  | 1,687 | 6.4% | 1,352 | 5.9% |  |
| 2008 | 1,266 | 7.0% | 373 | 5.1% |  | 1,797 | 6.8% | 1,260 | 5.5% |  |
| 2009 | 1,339 | 7.4% | 238 | 3.3% |  | 1,786 | 6.8% | 1,587 | 7.0% |  |
| 2010 | 1,382 | 7.6% | 263 | 3.6% |  | 1,634 | 6.2% | 944 | 4.1% |  |
| 2011 | 1,322 | 7.3% | 250 | 3.4% |  | 1,573 | 6.0% | 1,085 | 4.8% |  |
| 2012 | 1,120 | 6.2% | 347 | 4.8% |  | 1,465 | 5.6% | 1,094 | 4.8% |  |
| 2013 | 895 | 4.9% | 467 | 6.4% |  | 1,473 | 5.6% | 1,368 | 6.0% |  |
| 2014 | 935 | 5.1% | 455 | 6.3% |  | 1,431 | 5.4% | 1,288 | 5.7% |  |
| 2015 | 903 | 5.0% | 421 | 5.8% |  | 1,294 | 4.9% | 941 | 4.1% |  |
| 2016 | 1,057 | 5.8% | 535 | 7.4% |  | 1,532 | 5.8% | 1,333 | 5.9% |  |
| Hospital Type |  |  |  |  |  |  |  |  |  |  |
| Metropolitan | 14,157 | 77.8% | 5,396 | 74.2% | <0.00 | 20,493 | 77.8% | 16,942 | 74.4% | 0.17 |
| Urban | 4,037 | 22.2% | 1,879 | 25.8% |  | 5,846 | 22.2% | 5,836 | 25.6% |  |
| Clearance Time Category¶ |  |  |  |  |  |  |  |  |  |  |
| 1 Week | 14,368 | 79.0% | 3,052 | 42.0% | <0.00 | 17,370 | 65.9% | 14,663 | 64.4% | 0.67 |
| 2 Weeks | 2,383 | 13.1% | 1,910 | 26.3% |  | 4,840 | 18.4% | 4,329 | 19.0% |  |
| 3 or More Weeks | 1,443 | 7.9% | 2,313 | 31.8% |  | 4,129 | 15.7% | 3,786 | 16.6% |  |

| Neighborhood Income Decile |  |  |  |  |  |  |  |  |  |  |
| --- | --- | --- | --- | --- | --- | --- | --- | --- | --- | --- |
| Lowest Decile | 1,950 | 10.7% | 707 | 9.7% | 0.22 | 2,659 | 10.1% | 2,421 | 10.6% | 0.87 |
| 2nd Decile | 1,916 | 10.5% | 756 | 10.4% |  | 2,809 | 10.7% | 2,077 | 9.1% |  |
| 3rd Decile | 1,813 | 10.0% | 742 | 10.2% |  | 2,733 | 10.4% | 2,413 | 10.6% |  |
| 4th Decile | 1,905 | 10.5% | 784 | 10.8% |  | 2,933 | 11.1% | 2,790 | 12.2% |  |
| 5th Decile | 1,774 | 9.8% | 759 | 10.4% |  | 2,371 | 9.0% | 2,031 | 8.9% |  |
| 6th Decile | 1,756 | 9.7% | 645 | 8.9% |  | 2,390 | 9.1% | 2,244 | 9.9% |  |
| 7th Decile | 1,697 | 9.3% | 694 | 9.5% |  | 2,456 | 9.3% | 2,095 | 9.2% |  |
| 8th Decile | 1,749 | 9.6% | 708 | 9.7% |  | 2,772 | 10.5% | 2,122 | 9.3% |  |
| 9th Decile | 1,692 | 9.3% | 706 | 9.7% |  | 2,414 | 9.2% | 1,951 | 8.6% |  |
| Highest Decile | 1,677 | 9.2% | 673 | 9.3% |  | 2,345 | 8.9% | 2,367 | 10.4% |  |
| Unknown | 265 | 1.5% | 101 | 1.4% |  | 456 | 1.7% | 266 | 1.2% |  |

\* At the time of revascularization.
† Ejection Fraction at the time of revascularization; if missing, at the time of diagnostic catheterization.
§ Excluding atrial fibrillation and atrial flutter.
¶ Clearance time is the hypothetical time within which the wait list would be cleared at maximum weekly service capacity if there were no new arrivals.

Prior to adjustment with inverse probability of treatment weights, the patients undergoing delayed CABG, compared to patients undergoing timely PCI, had higher proportions of triple vessel disease, left main disease, male sex, a BMI >30, and an ejection fraction ≤50%. The delayed CABG group also had significantly higher proportions of atrial fibrillation or atrial flutter, congestive heart failure, diabetes, hypertension, and renal disease, compared to timely PCI. The timely PCI group had higher proportions of double-vessel disease and Canadian Cardiovascular Society (CCS) Angina Class 4. Patients were treated primarily in metropolitan hospitals, regardless of study group. Of the patients who underwent delayed CABG, 8.5% received only a saphenous vein graft (SVG), 71.6% received a single arterial graft, 16.3% received a double arterial graft, and 3.4% received a triple arterial graft. Of the patients who underwent timely PCI, 48.1% received bare-metal stents (BMS), 4.5% received a combination of BMS and drug-eluting stents (DES) and 42.8% received only DES. The mean wait time for CABG was 76 days, with the median at 50 days and the 90^th^ percentile at 162 days (Supplemental Material).

As expected, patients in the timely PCI group had a lower probability of being selected for delayed CABG than did those in the CABG group (Supplementary Material). However, there is overlap amongst the study groups – all patients had a positive probability of being assigned to either CABG or PCI. This is consistent with results of similarly designed studies of CABG versus PCI published elsewhere.^18^ All factors listed in Table 2 were used in the propensity score model.

### Outcome data and main results

The unadjusted CIF plot for compositive CV outcomes is shown in Figure 2; the adjusted CIF plot for composite CV outcomes is shown in Figure 3. Unadjusted and adjusted CVO point estimates at three years, with 95% confidence intervals are reported in Table 3.

**Figure 2.**
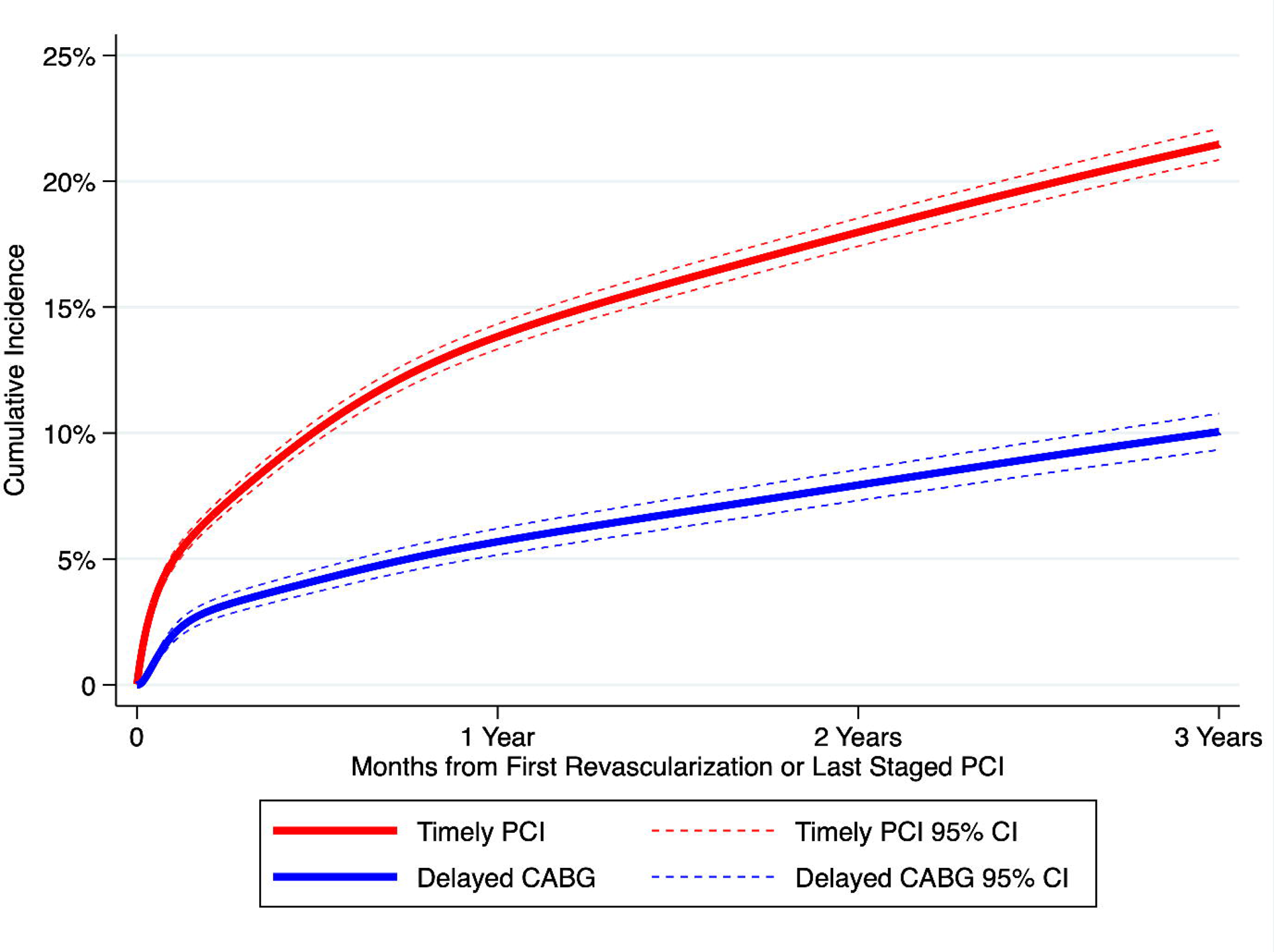
Unadjusted cumulative incidence functions with 95% confidence intervals for composite cardiovascular outcomes in the delayed CABG and timely PCI populations.

**Figure 3.**
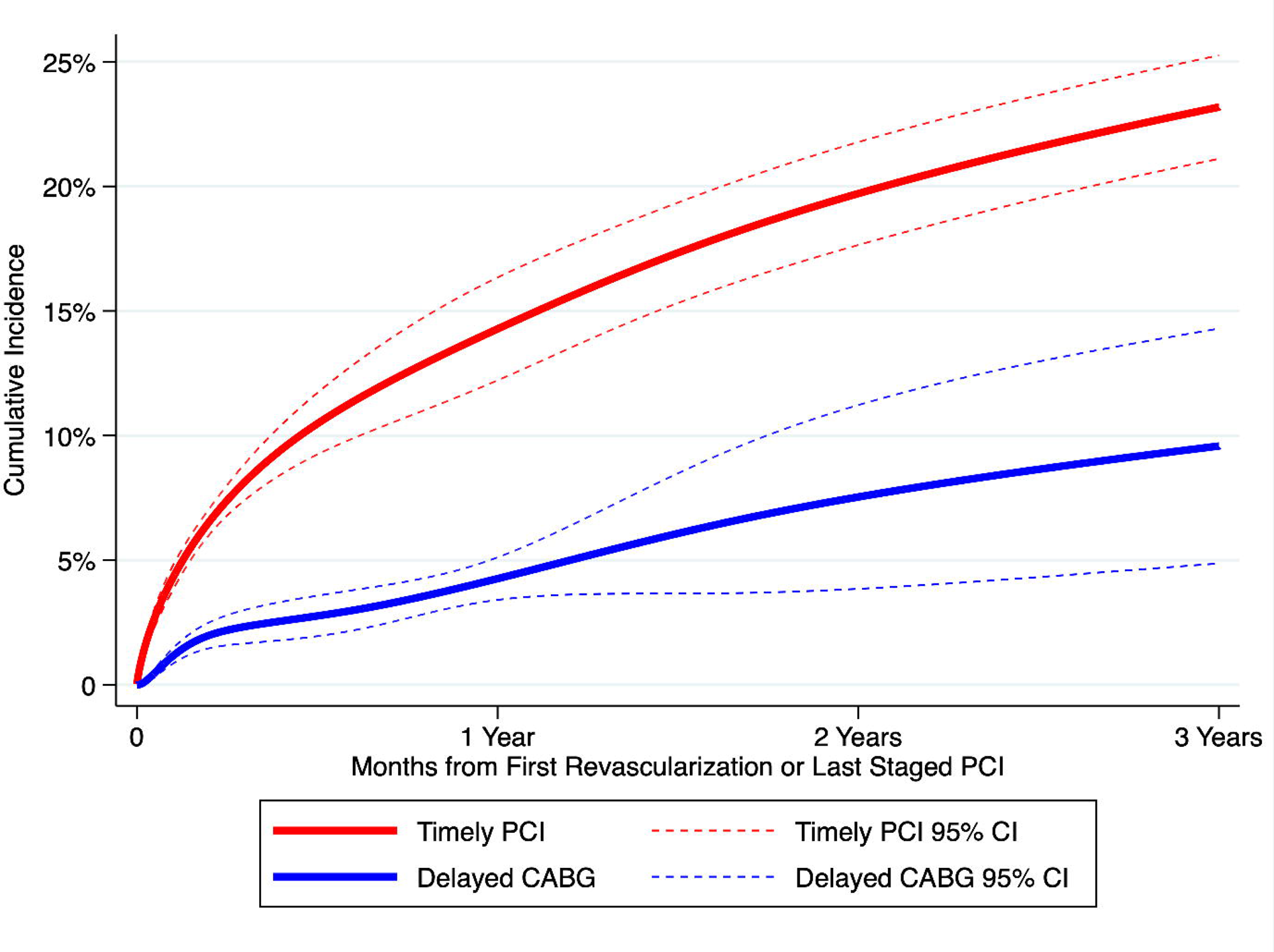
Adjusted cumulative incidence functions with 95% confidence intervals for composite cardiovascular outcomes in the delayed CABG and timely PCI populations.

**Table 3.** Cumulative incidence (percent) and 95% confidence intervals for composite cardiovascular outcomes in the delayed CABG and timely PCI populations, from unadjusted and adjusted analyses.

| **Unadjusted** | **30 Days** | **1 Year** | **2 Years** | **3 Years** |
| --- | --- | --- | --- | --- |
| Delayed CABG | 1.7 (1.4 – 1.7) | 5.7 (5.2 – 5.7) | 7.9 (7.3 – 7.9) | 10.1 (9.3 – 10.1) |
| Timely PCI | 4.5 (4.2 – 4.8) | 13.8 (13.3 – 14.3) | 18.0 (17.4 – 18.5) | 21.5 (20.8 – 22.1) |
| **Adjusted\*** |  |  |  |  |
| Delayed CABG | 1.0 (0.7, 1.0) | 4.3 (3.4 – 4.3) | 7.5 (3.9 – 7.5) | 9.6 (4.9 – 9.6) |
| Timely PCI | 3.8 (3.4 – 4.3) | 14.3 (12.2 – 16.3) | 19.7 (17.6 – 21.8) | 23.2 (21.1 – 25.3) |

\*Adjusted with inverse probability of treatment weights.

The 3-year unadjusted cumulative incidence of CVO was 10.1% in the delayed CABG group (95% confidence interval [CI], 9.3 – 10.8) and 21.5% (20.8 – 22.1) in the timely PCI group; the adjusted cumulative incidence was 9.6% in the delayed CABG group (95%, 4.8 – 14.4) compared to 23.2% in the timely PCI group (95% CI, 21.1 – 25.3). The unadjusted subdistribution hazard ratio of CVO for delayed CABG compared to timely PCI at three years was 0.55 (95% confidence interval [CI], 0.45 – 0.66); the adjusted subdistribution hazard was 0.50 (95% CI, 0.26 – 0.99), a lower hazard of CVO for delayed CABG compared to timely PCI. Unadjusted and adjusted CIF plots for mortality as a competing risk are shown in Figure 3. Unadjusted and adjusted cumulative mortality estimates, with 95% confidence intervals are shown in Table 4.

**Table 4.**
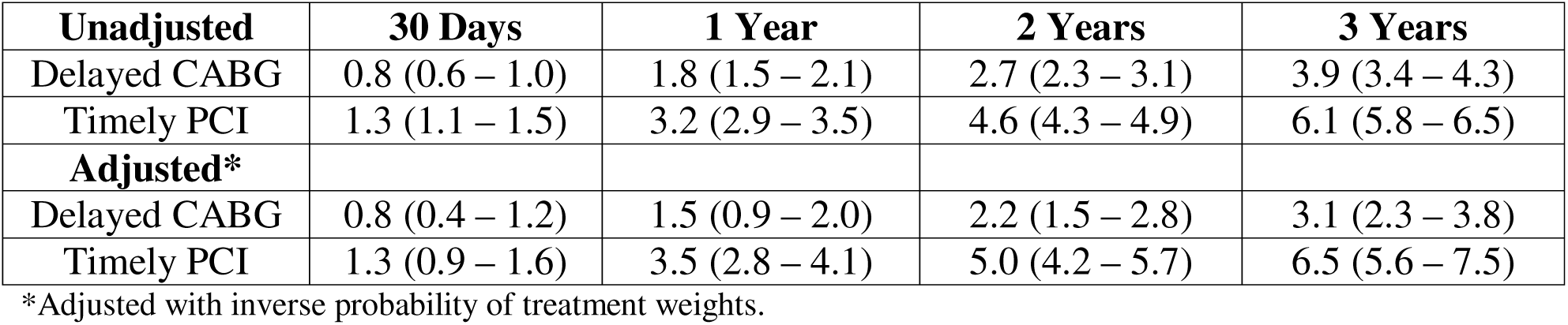
Cumulative incidence (percent) and 95% confidence intervals for mortality as a competing risk in the delayed CABG and timely PCI populations, from unadjusted and adjusted analyses.

| **Unadjusted** | **30 Days** | **1 Year** | **2 Years** | **3 Years** |
| --- | --- | --- | --- | --- |
| Delayed CABG | 0.8 (0.6 – 1.0) | 1.8 (1.5 – 2.1) | 2.7 (2.3 – 3.1) | 3.9 (3.4 – 4.3) |
| Timely PCI | 1.3 (1.1 – 1.5) | 3.2 (2.9 – 3.5) | 4.6 (4.3 – 4.9) | 6.1 (5.8 – 6.5) |
| **Adjusted\*** |  |  |  |  |
| Delayed CABG | 0.8 (0.4 – 1.2) | 1.5 (0.9 – 2.0) | 2.2 (1.5 – 2.8) | 3.1 (2.3 – 3.8) |
| Timely PCI | 1.3 (0.9 – 1.6) | 3.5 (2.8 – 4.1) | 5.0 (4.2 – 5.7) | 6.5 (5.6 – 7.5) |

\*Adjusted with inverse probability of treatment weights.

The 3-year unadjusted cumulative incidence of mortality was 3.9% in the delayed CABG group (95% CI 3.4 – 4.3) and 6.1% (95% CI, 5.8 – 6.5) in the timely PCI group; the adjusted cumulative incidence was 3.1% in the delayed CABG group (95% CI, 2.3 – 3.8) compared and 6.5% in the timely PCI group (95% CI, 5.6 – 7.5). The unadjusted subdistribution hazard ratio for mortality for delayed CABG compared to timely PCI at three years was 0.75 (95% CI, 0.56 – 1.01); the adjusted subdistribution hazard ratio at three years was 0.56 (95% CI, 0.32 – 0.98), a lower hazard for delayed CABG compared to timely PCI. It is important to interpret the competing risk results with caution to avoid the Table 2 fallacy^19^, as this analysis was not designed to estimate the cumulative incidence of mortality.

## DISCUSSION

### Key results

We drew on data from multiple population-based registries and databases to evaluate the effectiveness of delayed CABG as compared with timely PCI. In this study, we found that amongst British Columbia patients 60 aged years or older, who underwent non-emergency first-time revascularization for angiographically-proven, stable left main or multi-vessel ischemic heart disease in British Columbia, between January 1, 2001, and December 31, 2016, there was a significant difference in both unadjusted CVO and adjusted with inverse probability of treatment weights at and three years. The difference in CVO was established early, with the cumulative incidence function difference increasing throughout the remainder of the study period, starting to plateau after two years. We also found that a statistically significant difference in the cumulative incidence of mortality as a competing risk was established and sustained over the study period.

Our findings should be considered in the context of results from other studies. There have been eleven randomized controlled trials comparing CABG with PCI and stenting in patients with multi-vessel disease^20^ and six comparing CABG with PCI and stenting in specifically in patients with left-main disease.^21^ While none of these used the same operationalization of CVO as used in our study or include UA or CHF as outcomes, several reported unadjusted AMI and/or CVA outcomes. In patients with multi-vessel disease, the ARTS trial, which used BMS, included AMI as an outcome and found no significant differences between CABG and PCI in AMI at five years.^22^ Similarly, the ERACI-II trial found no statistically significant difference in AMI at five years^23^ The MASS-II trial reported no statistically significant difference in AMI at five years,^24^ but did find a significant difference at ten years.^25^ More recently, the SYNTAX trial, which used DES, found statistically significant lower rates of AMI in CABG patients at three years^26^ when compared to PCI patients using DES; this difference was sustained at five years.^27^ In the SYNTAX II study, comparing modern DES to the original SYNTAX CABG cohort, no significant difference was seen in AMI between CABG and PCI at two years^28^ In patients with left main disease, both the SYNTAX left main study^29^ and more recently, the NOBLE trial^30^ showed statistically significant differences in AMI favoring CABG at five years.

Several randomized controlled trials have reported CVA outcomes. In studies using BMS, no significant difference in CVA was observed in the ARTS-I^22^ or the MASS II trials at five years.^24^ The SYNTAX trial did observe a statistically significant difference in CVA at three years in favour of CABG.^26^ However, this was not observed at five years.^27^ SYNTAX II did show a statistically significant difference in CVA in favour of PCI, though the statistical finding was borderline and the number of events very small.^28^ In patients with left main disease, only SYNTAX reported a statistically significant difference at one year.^31^ Unfortunately, no randomized controlled trial of CABG versus PCI has reported frequencies of UA or CHF, either individually or as an element of a composite outcome, so no comparison with our results is available.

### Limitations

There are limitations to this study. First, it is possible that unmeasured confounders may affect our results. While we have successfully used inverse probability of treatment weights to balance differences in patient and health system factors in our study groups, these efforts may not be sufficient to fully account for between group differences. Second, our study period included only patients that underwent treatment between 2001 and 2016. Stent technology evolved significantly during that time,^32^ as did the use of antiplatelet therapy.^33^ We accounted for this by adjusting for calendar year of revascularization in the propensity score model. However, this may not have been adequate to account for these and any other potential confounders. Third, we make use of the DAD and the diagnosis codes it contains to operationalize our definitions of AMI, UA, CHF, and CVA. While validation of the results of abstracts have been completed, variation in coding practices amongst abstractors has been observed.^34^

### Interpretation

Our results suggest that there is evidence that patients in our study population who undergo delayed CABG experience a treatment benefit in cardiovascular disease progression over patients who undergo timely PCI. Further, these results suggest that the benefit is sustained over three years.

### Generalizability

Our results should be generalized to populations like that selected for this study and to health care systems that operate on a similar basis to that found in British Columbia. Physicians and policy makers should take caution in applying these results to other populations or health care systems.

## CONCLUSION

In summary, we collected data from the CSBC diagnostic catheterization, PCI, and CABG clinical registries, and linked them to the BC Ministry of Health DAD, the BC Vital Statistics Deaths File and Population Data BC’s Central Demographics File to assess the comparative effectiveness of timely PCI and delayed CABG on CVO in the presence of death as a competing risk. Patients older than 60 years of age with stable, multi-vessel or left-main ischemic heart disease that did not require emergency treatment had a significantly lower cumulative incidence and hazard of CVO and death as a competing risk when they received delayed CABG compared to timely PCI. Patients who must consider time to treatment, and physicians who must advise their patients and families, should be aware of these differences when selecting a revascularization strategy.

## DECLARATIONS

### ETHICS APPROVAL AND CONSENT TO PARTICIPATE

The University of British Columbia’s Clinical Research Ethics Board provided ethical approval of this research (Certificate Number H17-00505).

### CONSENT FOR PUBLICATION

The data stewards for this research provided their consent for publication.

### AVAILABILITY OF DATA AND MATERIALS

Access to data provided by the Data Stewards is subject to approval but can be requested for research projects through the Data Stewards or their designated service providers. All inferences, opinions, and conclusions drawn in this publication are those of the author(s), and do not reflect the opinions or policies of the Data Stewards.

### AUTHORS’ CONTRIBUTIONS

BS and GF conceived the original study concept. SH designed the study and acquired the data from the data stewards. SH designed the dataset creation plan (DCP) and the plan of analysis (POA). SH and LK created the analytical data set as established in the DCP. SH performed all statistical analyses in the POA. All authors contributed to the content of the DCP and POA. All authors contributed to the interpretation of the analyzed data. All authors critically revised the manuscript. All authors approved the final version for submission.

## ACKNOWLEDGEMENTS

We gratefully acknowledge the patients whose anonymized cardiac information is collected in the Cardiac Services BC database, without whom this work would not be possible.

## LIST OF ABBREVIATIONS

AHA: American Heart Association
AMI: Acute Myocardial Infarction
BMS: Bare Metal Stent
CABG: Coronary Artery Bypass Graft
CCS: Canadian Cardiovascular Society
CIHI: Canadian Institute for Health Information
CHF: Congestive Heart Failure
CIF: Cumulative Incidence Function
CSBC: Cardiac Services BC
CVA: Cerebrovascular Accident
CVO: Cardiovascular Outcomes
DAD: Discharge Abstract Database
DES: Drug-Eluting Stent
ICD-9: International Classification of Diseases, Ninth Revision
ICD-10-CA: International Classification of Diseases, Tenth Revision, Canada
PCI: Percutaneous Coronary Intervention
STROBE: Strengthening the Reporting of Observational Studies in Epidemiology
UA: Unstable Angina

