## Supplementary Material for "The effect of coronary revascularization timing on cardiovascular outcomes in patients with ischemic heart disease"

In this supplementary material file, we provide information on key concept operationalization used in this study, the algorithm used to identify repeat revascularization, the rule applied to identify staged Percutaneous Coronary Intervention (PCI), the propensity score distribution in the study groups, information on additional variables and the source data sets, as well as the STROBE Checklist for the standard reporting of observational studies.

1. Cardiovascular Outcomes Operationalization
2. Propensity Scores and Standardized Differences for Delayed Coronary Artery Bypass Grafting (CABG) in the Timely PCI and Delayed CABG Populations
3. Wait Time Distribution Median, Mean, and 90^th^ Percentile by Study Group
4. Comorbidity Operationalization
5. Staged PCI Identification Rule
6. Repeat Revascularization Algorithm
7. Additional Variables
8. STROBE Checklist for Cohort Studies
9. CSBC Data Set Background
10. Data Source Citations
11. References

**1. Cardiovascular Outcomes Operationalization**

Dates of cardiovascular outcomes - unstable angina, acute myocardial infarction (AMI), cerebrovascular accident (CVA), and congestive heart failure (CHF) – were identified from the first hospital admission after discharge from index revascularization or last staged PCI, with the specified diagnosis codes and types in the Discharge Abstract Database specified. These cardiovascular outcomes were identified using the following fields and values:

### Date

Date of the first hospital admission after discharge from index revascularization in ADDATE field.

### Diagnosis Codes

ICD-10-CA or ICD-9 diagnosis codes for unstable angina, AMI, CVA, or CHF (Table 1) in DIAGX1 – DIAGX25 or DIAG1 – DIAG16 fields, respectively.

### Diagnosis Types

Values ‘M’, ‘1’, ‘2’, ‘6’, ‘W’, ‘X’, or ‘Y’ in DTYPX1 – DTYPX25 fields or DTYP1 – DTYP16 fields. These values represent the most responsible diagnosis, pre-admit comorbidity, post-admit comorbidity, proxy most responsible diagnosis, and the first, second, and third service transfers.

Table A1. ICD-10-CA and ICD-9 codes used to identify cardiovascular outcomes.

| **Cardiovascular Outcome** | **ICD-10-CA Codes** | **ICD-9 Codes** |
| --- | --- | --- |
| Unstable Angina | I20.0 | 411.^ |
| AMI | I21.^ | 410.^ |
|  | I22.^ | 410.^ |
| CVA | I60.^ | 430 |
|  | I61.^ | 431 |
|  | I62.9 | 432.9 |
|  | I63.0, I63.1, I63.2 | 433.9 |
|  | I63.3 | 434.0 |
|  | I63.4 | 434.1 |
|  | I63.5 | 434.9 |
|  | I63.8 | 434.9 |
|  | I63.9 | 434.9 |
|  | H34.1 | 362.3 |
|  | I64 | 436 |
|  | I63.6 | 437.6 |
|  | I67.6 | 437.6 |
|  | G08 | 325 |
| CHF | I50.^ | 428.^ |

Codes appended with the carat symbol (‘^’) will include all subcodes in the search for diagnosis codes.

**2. Propensity Scores and Standardized Differences for Delayed Coronary Artery Bypass Grafting (CABG) in the Timely PCI and Delayed CABG Populations**

Figure A1. Propensity scores for delayed Coronary Artery Bypass Grafting (CABG) in the timely Percutaneous Coronary Intervention and delayed CABG populations.

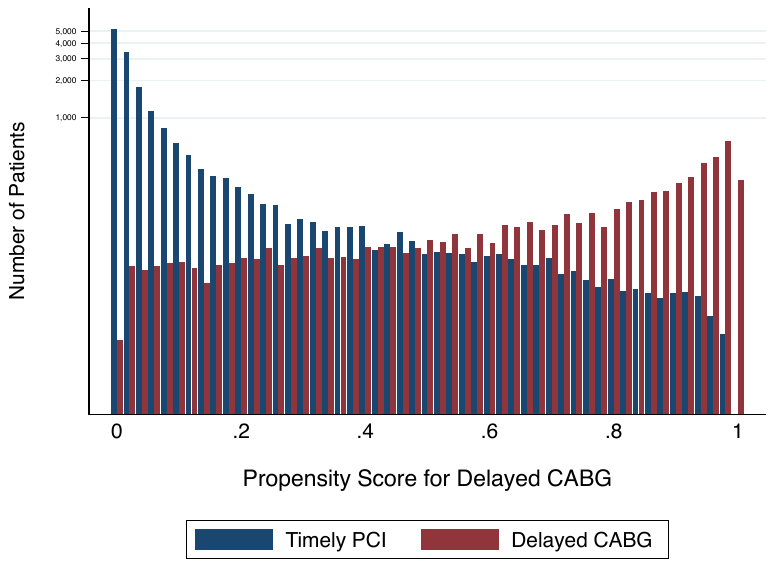

Figure A2.

Standardized differences between study groups in propensity score model factors before and after inverse probability of treatment weighting.

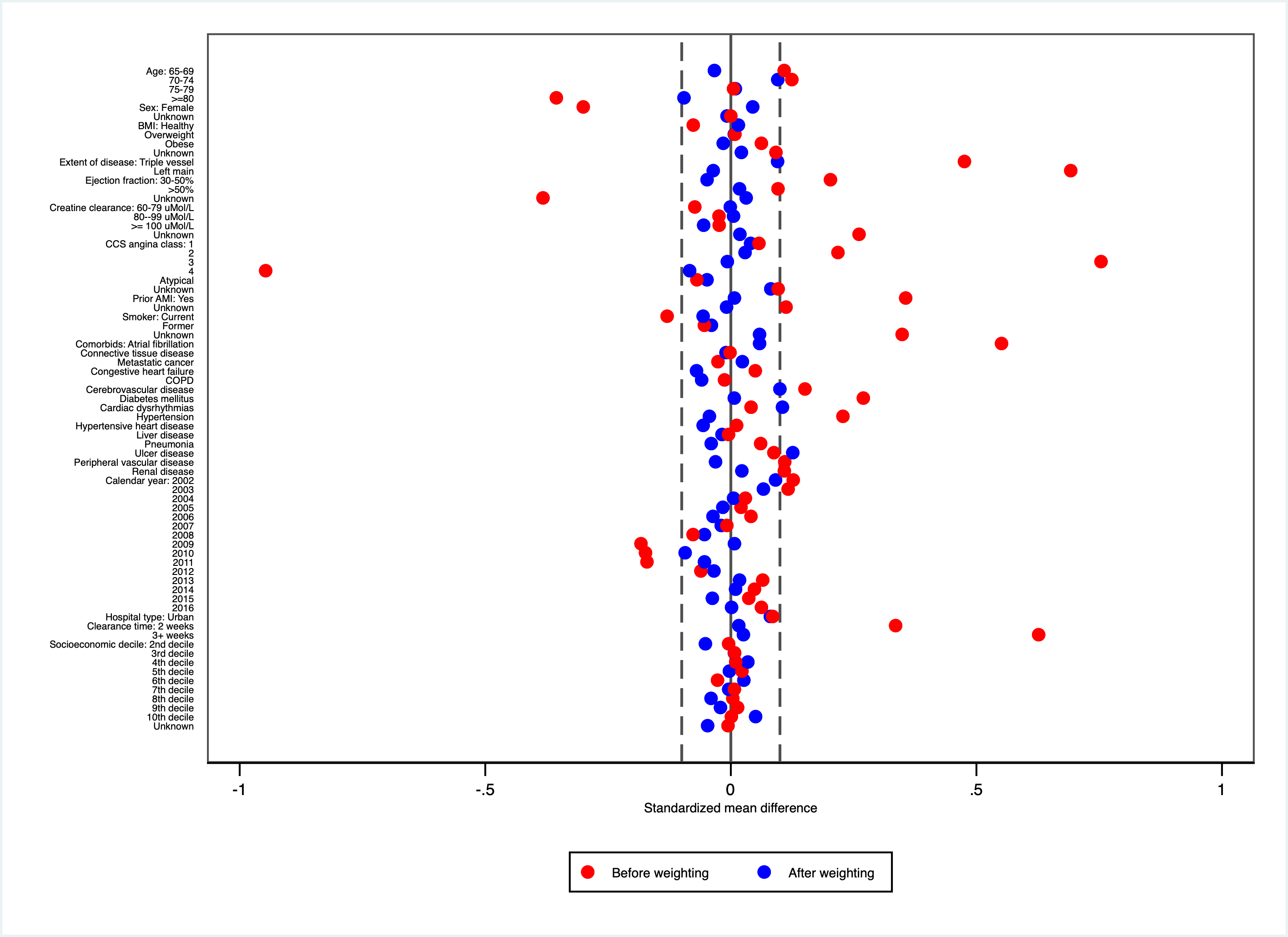

**3. Wait Time Distribution Median, Mean, and 90^th^ Percentile by Study Group**

Table A2.

Median, mean, and 90th percentile wait times by study group, in days.

| **Study Group** | **Mean** | **50^th^ Percentile** | **90^th^ Percentile** |
| --- | --- | --- | --- |
| Timely PCI | 16 Days | 3 Days | 39 Days |
| Delayed CABG | 76 Days | 50 Days | 162 Days |

Reasons for CABG delay were not consistently available across the duration of the study period.

**4. Clearance Time Operationalization**

*Background*

Clearance time is the hypothetical time within which the list will be cleared at maximum capacity if there are no new arrivals.^1^

For each patient, we estimated the clearance time by dividing the number of pre-surgical or pre-catheterization patients present on the waiting list on the day a patient is booked for their procedure by the maximum weekly procedure rate at the same hospital.

We used the waiting list calculation proposed by Cottrell^2^ and stratified the waiting list into four strata as used by Sobolev et al.^1^ We categorize dthe list size as a function of clearance time. We divided the list into four wait list categories: (1) where capacity is adequate to meet demand (2) where there is a short clearance time, or (3) where there is a prolonged clearance time. We used weekly clearance time as the reference value.

Table A3

Clearance Times Used to Estimating Demand for Coronary Revascularization Procedures

| **Category** | **Category Label** | **Clearance Time** |
| --- | --- | --- |
| 1 | 1 Week | <1 Week |
| 2 | 2 Weeks | ≥ 1 Week to  <2 Weeks |
| 3 | 3 or More Weeks | ≥ 2 Weeks |

Originally, we established four wait list categories characterizing clearance time, based on the recommendations made by the Canadian Cardiovascular Society’s Access to Care Working Group^3^ for access to cardiac surgery and cardiac catheterization services. For both procedures, access is recommended within two weeks for urgent patients and six weeks for all other patients. However, initial implementation suggested that a small number of CABG patients had a clearance time greater than six weeks, and no PCI patients had a clearance time of greater than six weeks. Therefore, the few CABG patients in the greater than six-week category were combined with the greater than two-week category to provide for meaningful categorizations.

*Clearance Time Calculation*

For each patient, we estimated the clearance time by dividing the number of pre-surgical or pre-catheterization patients present on the waiting list on the day a patient was booked for their procedure by the maximum weekly procedure rate at the same hospital.

We counted the number of patients present on the waiting list based on (1) the existing list size at the time the patient is booked and (2) the batch size of new arrivals.^1^ For each patient, the list size was a census of patients with equal or higher priority present at the date of booking in one of the cardiac centres. Patients contributed one count to the list size for each week on the list, except for the week of arrival. Patients who underwent revascularization were considered removed from the waitlist in the week prior to their procedure, as patients were scheduled on a weekly basis. For each patient, the batch size was the count of patients with higher or equal priority registered on the list in the same week.

The maximum rate estimated the largest number of cardiac surgeries or cardiac catheterizations a hospital could deliver during a week, thereby providing means to estimate the hospital’s capacity to manage existing demand.

**Clearance Time Dependencies**

Clearance time depends on several features of the revascularization system.

Table A4. Maximum Weekly Procedure Rates for Cardiac Surgery and Cardiac Catheterization Lab Procedures at British Columbia Cardiac Centres

| **Cardiac Centre** | **Weekly Cardiac Surgery Procedures** | **Weekly Cardiac Catheterization Lab Procedures** |
| --- | --- | --- |
| Kelowna General Hospital | 12 | 60 |
| Royal Columbian Hospital | 20 | 88 |
| Royal Jubilee Hospital | 15 | 60 |
| St. Paul’s Hospital | 19 | 55 |
| Vancouver General Hospital | 16 | 75 |

Table A4 summarizes select features of the coronary revascularization system relevant to the estimation of clearance time. We obtained the weekly cardiac surgery and weekly cardiac catheterization lab procedure volumes from Cardiac Services BC (CSBC), who sought data from the triage coordinators for each site and service. These figures constitute site capacity in our operationalization of demand. We validated these figures as representative of the study period through personal communication with cardiac surgery division heads and site interventional cardiology medical directors for each site.

*Clearance Time by Revascularization Procedure*

CABG and PCI clearance times are different procedures, with different resource requirements. We accounted for these differences by separating the calculations for cardiac catheterization procedures and cardiac operating room procedures.

While the calculation of clearance time is for cardiac surgery requires only counting cardiac surgery cases, the cardiac catheterization lab procedures require counting both diagnostic catheterization cases and scheduled PCI cases as a procedure slot can be used for both procedures. We counted diagnostic catheterization cases, ad-hoc PCI cases, and scheduled PCI cases in this category, as each consumes a cardiac catheterization procedure slot.

*Clearance Time and Booking Hospital*

CABG and PCI clearance times are dependent on hospital capacity. Therefore, we accounted for differences using capacity estimates obtained from the cardiac centres.

If a patient record had an unknown booking hospital, we used instead the hospital that performs the revascularization as the booking hospital. If a patient record had a booking hospital where only diagnostic catheterizations are performed, specifically Lion’s Gate Hospital, then we attributed demand to the revascularizing hospital.

*Clearance Time and Booking Priority*

CABG and PCI clearance times are dependent on booking priority. Therefore, we accounted for booking priority in the clearance time calculation.

If a patient record had an unknown priority at time of booking, we used the priority at the time of revascularization.

*Clearance Time and Calendar Week of Booking & Procedure*

CABG and PCI clearance times are dependent on the calendar week of the booking procedure. Therefore, we accounted for calendar week in the batch size calculation used in estimating clearance time.

If a patient had an unknown booking date, we used the date of diagnostic catheterization as the proxy booking date. If a patient had an unknown booking date for the diagnostic catheterization, we assumed that the booking happened the same week of the procedure.

**4. Comorbidity Operationalization**

We used linked care episodes with hospitalization records in the Discharge Abstract Database to identify comorbidities. To identify comorbidities, we searched for diagnosis codes between the date of revascularization and the date that is one year prior to the date of the diagnostic catheterization.

Comorbidity codes were identified through previous work completed by members of the study team, and informed by clinical advisor feedback, and the Canadian Institute for Health Information (CIHI) and Canadian Cardiovascular Society’s (CCS) Cardiac Care Quality Indicators (CCQI) initiative methodology.

Table A5. ICD-10-CA and ICD-9 codes used to identify comorbidities.

| **Comorbid condition** | **ICD-10-CA code** | **ICD-9 code** |
| --- | --- | --- |
| Any Tumor or Metastatic Solid Tumor* | C00*-C75*;  C77*;  C78*;  C79*;  C80*; | 140 – 195;  196;  197;  198;  199; |
| Atrial Fibrillation or Atrial Flutter | I48.*; | 427.3; |
| Cardiac Dysrhythmias Excluding Atrial Fibrillation and Atrial Flutter | I47.*;  I49.* | 427.8, 427.0, 427.1, 427.2;  427.4, 427.6, 427.8, 427.9 |
| Cerebrovascular Disease | I60*;  I61*;  I62*;  I63*;  I64*;  I65*;  I67*;  I68*;  G45*;  G81*; | 430;  431;  432.1, 432.0, 432.9;  433.9, 434.0, 434.1, 434.9, 437.6;  436;  433.2, 433.0, 433.1, 433.3, 433.8. 433.9;  437.8, 437.3, 437.0, 437.2, 437.5, 437.6, 437.4, 437.9;  437.8, 437.4, 437.8;  435, 362.3;  342.0, 342.1, 342.9; |
| Chronic Pulmonary Disease | J40*;  J41*;  J42*;  J43*;  J44*;  J45*;  J47* | 490;  491.0, 491.1, 491.8;  491.9;  492;  496, 491.2;  493.0, 493.1, 493.9;  494; |
| Congestive Heart Failure | I50* | 428 |
| Connective Tissue Disease | M30*;    M31*,  M32*,    M33*,  M34*,  M35*,  M36*,  M05*,  M06*,  M07*,  M08*,  M09*,  M10*,  M11*,  M12*,  M13*,    M14* | 446.0, 446.4, 447.8, 446.1;  446.2, 446.6, 446.3, 446.4, 446.7. 446.5, 446.0, 447.8, 447.5;  710.0;  710.3, 710.4;  710.1;  710.2, 710.8, 136.1, 725, 729.4, 710.8, 729.3, 728.5, 710.8, 710.9;  710.3, 713.2, 713.6, 713.0;  714.1, 714.8, 714.2, 714.0;  714.0;  713.3, 713.1;  714.3, 720.0, 714.3;  714.3;  274.0. 984.9, 274.9, 274.8;  275.4, 712.8, 712.9;  714.4, 716.0, 719.2, 719.3, 716.1, 716.8;  716.5, 716.6, 716.8, 716.9;  713.0, 713.3, 713,7, 713.5, 713.8 |
| Dementia | F00*;  F01*;  F02*;  F03*;  F10*; | 290.1;  290.0;  290.8;  No value;  305.0, 303, 291.8, 291.3, 291.1, 291.2, 291.9; |
| Diabetes | E10*;  E11*;  E13*;  E14*; | 250.2, 250.1, 250.3, 250.4; 250.5, 250.6, 250.7, 250.0;  250.2, 250.1, 250.3, 250.4,  250.5, 250.6, 250.7, 250.0;  250.1, 250.3, 250.4, 250.5, 250.6, 250.7, 250.0; |
| Endocarditis | I01.1;  I33.*;    I38.*;  I39.8; | 391.1;  421.0, 421.9;  424.9;  424.9; |
| Hypertension | I10.*;  I12.*;  I13.*;  I15.*; | 401.1, 401.0;  403.9;  404.9;  405.9, 405.0; |
| Hypertensive heart disease, other rheumatic heart diseases | I09*,  I11* | 398.0, 397.9, 393, 398.9;  402.9; |
| Leukemia* | C91*;  C92*;  C93*;  C94*;  C95*;  C96* | 204.0, 204.1, 204.9, 202.4, 204.8, 204.9;  205.0, 205.1, 205.3, 205.8, 205.9;  206.0, 206.8, 206.9;  207.0, 207.2, 207.8, 289.8, 207.8;  208.0, 208.1, 208.8, 208.9;  202.5, 202.3, 202.6, 200.0, 202.8, 202.9; |
| Liver Disease (Mild or Moderate or Severe or Viral) | K70*;  K71*;  K72*;  K73*,  K74*,  K75*,  K76*,  K77*;  B15*;    B16*;  B17*;    B18*;  B19*; | 571.0, 571.1, 571.2, 572.8, 571.3;  573.3;  570, 572.8;  571.4;  571.5, 571.9, 571.6;  572.0, 572.1, 572.3;  571.8, 573.0, 570, 573.4, 573.8, 572.3, 572.4, 573.8, 573.9;  573.2, 572.8;  070.0, 070.1;  070.2, 070.3;  070.5, 070.9;  070.3. 070.5, 070.9;  070.6, 070.9; |
| Lymphoma* | C81*;  C82*;    C83*;  C84*;    C85*;  C86*;    C88*;    C90*; | 201.4, 201.5, 201.6, 201.7, 201.1, 201.9;  202.0, 202.8  202.8, 200.8, 200.1, 200.2;  202.1, 202.2, 202.0;  202.8;  No Code;  273.3, 200.8, 203.8;  203.0; |
| Peripheral Vascular Disease | I70;    I71*;  I72*;  I73*;  I74*;  I77*;  I78*;  I79*;  M30*; | 440.0, 440.1, 440.2, 440.8, 440.9;  441.0, 441.1, 441.2, 441.3, 441.4, 441.5, 441.5, 441.6;  442.8, 442.0, 442.1, 442.2, 442.3, 442.8, 442.9;  443.0, 443.1, 443.8, 443.9;  444.0, 444.1, 444.2, 444.8, 444.9;  447.0, 447.1, 447.2, 447.8, 447.4, 447.5, 447.6, 447.8, 447.9;  448.0, 448.1, 448.9;  441.7, 447.7, 443.8, 448.9;  446.0; |
| Pneumonia | J10.0;  J11.0;  J12.*;  J13;  J14;  J15.*;  J16.8;  J18.*;  J85.1; | 487.0;  487.0;  480.0, 480.1, 480.2, 480.8, 480.9;  481;  482.2;  482.0, 482.1, 482.4, 482.3, 482.8, 483, 482.9;  483;  485, 481, 514, 486;  513.0; |
| Renal Disease (Moderate or Severe) | I12*;  I13*;  N00*;  N01*;  N02*;  N03*;  N04*;  N05*;  N17*;  N18*;  N19*; | 403.9;  404.9;  580.8. 580.0, 580.9;  583.4;  599.7;  582.8, 582.1, 582.0, 582.2, 582.9;  581.3, 581.1, 581.0, 581.2, 581.8, 581.9;  581.3, 583.8, 583.1, 583.0, 583.2, 583.9;  584.5, 584.6, 584.7, 584.8, 584.9;  585;  586; |
| Ulcer Disease | K25*;  K26*;    K27*,  K28* | 531.0, 531.1, 531.2, 531.3, 531.4, 531.5, 531.6, 531.7, 531.9;  532.0, 532.1, 532.2, 532.3, 532.4, 532.5, 532.6, 532.7, 532.9;  533.0, 533.1, 533.2, 533.3, 533.4, 533.5, 533.5 533.6, 533.7, 533.9;  534.0, 534.1, 534.2, 534.3, 534.4, 534.5, 534.6, 534.7, 534.9; |

* The *Any Tumor or Metastatic Tumor*, *Leukemia*, and *Lymphoma* variables were merged in the analytical data set into a single variable, *Metastatic Cancer*.

**6. Staged PCI Identification Rule**

Patients who undergo staged PCI do not complete their planned course of treatment at index revascularization. Cardiac Services BC data on staged PCI was incomplete, identifying only seven patients undergoing staged PCI between 2005 and 2011. Therefore, a rule was needed to identify patients with multiple PCI records to differentiate them from patients with repeat revascularization.

Patients who undergo staged PCI do not complete their planned course of treatment at index revascularization. For patients with multiple PCI records having subsequent diagnostic catheterization dates the same or within one Sunday of the one at index, then the date revascularization is achieved is the date of the last PCI if it falls within 60 days of index PCI and 30 days of the non-index PCI. For any unresolved records, we review each record to determine if a last staged PCI date can be identified using the diagnostic catheterization date, the PCI date, and the event type. Any records remaining after the review are considered unresolved and are set aside.

**7. Repeat Revascularization Algorithm**

Repeat revascularization will be identified by the first CABG or PCI occurring after live discharge from index revascularization, when a corresponding DC date differs from the DC date for the index revascularization. In addition, repeat revascularization with PCI will follow index PCI after 60 days or non-index PCI after 30 days. Repeat revascularization is identified using CSBC records.

**8. Additional Variables**

We used variables in the form in which they were received from the data stewards. Some concepts, such as comorbidities and clearance time, were operationalized by the study team from data already in the data set. Comorbidities were identified from diagnosis codes in the DAD, using work completed in previous research by the study investigators^1,4^ as a foundation, and informed by clinical advisor feedback and the CIHI/CCS Cardiac Care Quality Initiative’s comorbidity methodology.^5^ Body mass index was derived from height and weight data in the CSBC registry and then categorized to address extreme values found in a small number of records. CABG patients whose CCS angina grade^6^ was classified as Class 4A, 4B, or 4C in the CSBC cardiac surgery registry were grouped to Class 4. Clearance time was defined as the hypothetical time within which the wait list would be cleared at maximum weekly service capacity if there were no new arrivals.^2,7^

**9. STROBE Checklist for Cohort Studies**

|  | Item # | Recommendation |  | Reported On Page # |
| --- | --- | --- | --- | --- |
| **Title and abstract** | 1 | (*a*) Indicate the study’s design with a commonly used term in the title or the abstract |  | 1 |
|  |  | (*b*) Provide in the abstract an informative and balanced summary of what was done and what was found |  | 2 |
| Introduction | | |  |  |
| Background/rationale | 2 | Explain the scientific background and rationale for the investigation being reported |  | 3 |
| Objectives | 3 | State specific objectives, including any prespecified hypotheses |  | 3 |
| Methods | | |  |  |
| Study design | 4 | Present key elements of study design early in the paper |  | 3 |
| Setting | 5 | Describe the setting, locations, and relevant dates, including periods of recruitment, exposure, follow-up, and data collection |  | 5 |
| Participants | 6 | (*a*) Give the eligibility criteria, and the sources and methods of selection of participants. Describe methods of follow-up |  | 4 |
|  |  | (*b*) For matched studies, give matching criteria and number of exposed and unexposed |  | 4, 5 |
| Variables | 7 | Clearly define all outcomes, exposures, predictors, potential confounders, and effect modifiers. Give diagnostic criteria, if applicable |  | 4, 5 |
| Data sources/ measurement | 8* | For each variable of interest, give sources of data and details of methods of assessment (measurement). Describe comparability of assessment methods if there is more than one group |  | 4 |
| Bias | 9 | Describe any efforts to address potential sources of bias |  | 5, 6 |
| Study size | 10 | Explain how the study size was arrived at |  |  |
| Quantitative variables | 11 | Explain how quantitative variables were handled in the analyses. If applicable, describe which groupings were chosen and why |  | 7, 8 |
| Statistical methods | 12 | (*a*) Describe all statistical methods, including those used to control for confounding |  | 6, 7 |
|  |  | (*b*) Describe any methods used to examine subgroups and interactions |  | N/A |
|  |  | (*c*) Explain how missing data were addressed |  | 6 |
|  |  | (*d*) If applicable, explain how loss to follow-up was addressed |  | N/A |
|  |  | (*e*) Describe any sensitivity analyses |  | N/A |
| Results | | |  |  |
| Participants | 13* | (a) Report numbers of individuals at each stage of study—eg numbers potentially eligible, examined for eligibility, confirmed eligible, included in the study, completing follow-up, and analysed |  | 7 |
|  |  | (b) Give reasons for non-participation at each stage |  | Figure 1 |
|  |  | (c) Consider use of a flow diagram |  | Figure 1 |
| Descriptive data | 14* | (a) Give characteristics of study participants (eg demographic, clinical, social) and information on exposures and potential confounders |  | 7  Table 2 |
|  |  | (b) Indicate number of participants with missing data for each variable of interest |  | Table 2 |
|  |  | (c) Summarise follow-up time (eg, average and total amount) |  | Figure 2, 3 |
| Outcome data | 15* | Report numbers of outcome events or summary measures over time |  | 7, 8 |
| Main results | 16 | (*a*) Give unadjusted estimates and, if applicable, confounder-adjusted estimates and their precision (eg, 95% confidence interval). Make clear which confounders were adjusted for and why they were included |  | 8 |
|  |  | (*b*) Report category boundaries when continuous variables were categorized |  | Table 2 |
|  |  | (*c*) If relevant, consider translating estimates of relative risk into absolute risk for a meaningful time period |  | N/A |
| Other analyses | 17 | Report other analyses done—eg analyses of subgroups and interactions, and sensitivity analyses |  | N/A |
| Discussion | | |  |  |
| Key results | 18 | Summarise key results with reference to study objectives |  | 8 |
| Limitations | 19 | Discuss limitations of the study, taking into account sources of potential bias or imprecision. Discuss both direction and magnitude of any potential bias |  | 10 |
| Interpretation | 20 | Give a cautious overall interpretation of results considering objectives, limitations, multiplicity of analyses, results from similar studies, and other relevant evidence |  | 10 |
| Generalisability | 21 | Discuss the generalisability (external validity) of the study results |  | 10 |
| Other information | | |  |  |
| Funding | 22 | Give the source of funding and the role of the funders for the present study and, if applicable, for the original study on which the present article is based |  | 12 |

**10. CSBC Data Set Background**

The CSBC database is a prospective registry of cardiovascular procedures. CSBC, a program of the Provincial Health Services Authority, has province-wide responsibility for planning, funding, and evaluating cardiovascular services in British Columbia, a western Canadian province serving approximately five million residents. All coronary revascularization procedures are recorded and stored in the database. Reporting is mandatory, subject to local audit processes, generates operative reports and assists billing. As such, procedural capture is considered near complete. Coronary revascularization occurs across five publicly funded centres. Access to care is universal and based solely on physician assessment of need rather than ability to pay. All patients are entered into the database without requirement for informed consent, therefore avoiding participation bias.

**11. Data Source Citations**

Access to data provided by the Data Steward(s) is subject to approval but can be requested for research projects through the Data Steward(s) or their designated service providers. All inferences, opinions, and conclusions drawn in this publication are those of the author(s), and do not reflect the opinions or policies of the Data Steward(s)

Cardiac Services BC [creator] (2016): Cardiac Services BC Cardiac Registries. Cardiac Services BC [publisher]. Data Extract. CSBC (2018).

Canadian Institute for Health Information [creator] (2018): Discharge Abstract Database (Hospital Separations). V2. Population Data BC [publisher]. Data Extract. MOH (2018).

British Columbia Ministry of Health [creator] (2018): Vital Events Deaths. V2. Population Data BC [publisher]. Data Extract. MOH (2018).

British Columbia Ministry of Health [creator] (2018): Consolidation File (MSP Registration & Premium Billing). V2. Population Data BC [publisher]. Data Extract. MOH (2018).
